# Differential Peripheral Blood Glycoprotein Profiles in Symptomatic and Asymptomatic COVID-19

**DOI:** 10.1101/2022.01.07.21267956

**Authors:** Chad Pickering, Bo Zhou, Gege Xu, Rachel Rice, Prasanna Ramachandran, Hector Huang, Tho D. Pham, Jeffrey M. Schapiro, Xin Cong, Saborni Chakraborty, Karlie Edwards, Srinivasa T. Reddy, Faheem Guirgis, Taia T. Wang, Daniel Serie, Klaus Lindpaintner

**Affiliations:** InterVenn Biosciences, South San Francisco, CA; University of Florida, Jacksonville, FL; Department of Medicine, Division of Infectious Diseases, Stanford University, Stanford, CA; Blood Center, Palo Alto, CA; TPMG Regional Reference Laboratory, Kaiser Permanente Northern California, Berkeley, CA; University of California Los Angeles, Department of Molecular and Medical Pharmacology, Los Angeles, CA

**Keywords:** COVID-19, SARS-CoV-2, glycoproteomics, biomarkers, glycosylation

## Abstract

Glycosylation is the most common form of post-translational modification of proteins, critically affecting their structure and function. Using liquid chromatography and mass spectrometry for high-resolution site-specific quantification of glycopeptides coupled with high-throughput artificial intelligence-powered data processing, we analyzed differential protein glyco-isoform distributions of 597 abundant serum glycopeptides and non-glycosylated peptides in 50 individuals who had been seriously ill with COVID-19 and in 22 individuals who had recovered after an asymptomatic course of COVID-19. As additional comparison reference phenotypes, we included 12 individuals with a history of infection with a common cold coronavirus, 16 patients with bacterial sepsis, and 15 healthy subjects without history of coronavirus exposure. We found statistically significant differences, at FDR<0.05, for normalized abundances of 374 of the 597 peptides and glycopeptides interrogated, between symptomatic and asymptomatic COVID-19 patients. Similar statistically significant differences were seen when comparing symptomatic COVID-19 patients to healthy controls (350 differentially abundant peptides and glycopeptides) and common cold coronavirus seropositive subjects (353 differentially abundant peptides and glycopeptides). Among healthy controls and sepsis patients, 326 peptides and glycopeptides were found to be differentially abundant, of which 277 overlapped with biomarkers that showed differential expression between symptomatic COVID-19 cases and healthy controls. Among symptomatic COVID-19 cases and sepsis patients, 101 glycopeptide and peptide biomarkers were found to be statistically significantly abundant. Using both supervised and unsupervised machine learning techniques, we found specific glycoprotein profiles to be strongly predictive of symptomatic COVID-19 infection. LASSO-regularized multivariable logistic regression and K-means clustering yielded accuracies of 100% in an independent test set and of 96% overall, respectively. Our findings are consistent with the interpretation that a majority of glycoprotein modifications observed which are shared among symptomatic COVID-19 and sepsis patients likely represent a generic consequence of a severe systemic immune and inflammatory state. However, there are glyco-isoform changes that are specific and particular to severe COVID-19 infection. These may be representative of either COVID-19-specific consequences or of susceptibility to or predisposition for a severe course of the disease. Our findings support the potential value of glycoproteomic biomarkers in the biomedical understanding, and, potentially, the clinical management of serious acute infectious conditions.

## 1. Introduction

Coronavirus disease 2019 (COVID-19) is a highly contagious infectious disease caused by the severe acute respiratory syndrome coronavirus 2 (SARS-CoV-2). The illness, characterized in severe cases by respiratory distress syndrome, was initially recognized in December 2019 in the city of Wuhan, Peoples Republic of China, spreading subsequently across the country and, very quickly, across the world as a pandemic of unprecedented impact and duration. As of November 2021, the pandemic of COVID-19 has affected more than 260 million individuals. Although the majority of COVID-19 cases generally only suffer mild symptoms, or remain fully asymptomatic, the pandemic has caused more than 5.1 million deaths worldwide, with many more having experienced a serious and life-threatening illness. Many studies have been conducted to identify characteristics and potential clinical, demographic, and epidemiological risk factors of becoming seriously ill with COVID-19 infection. Diabetes mellitus, cardiovascular diseases, hypertension, chronic respiratory diseases, as well as advanced age, male sex, sociocultural factors, and ethnicity have all been found to be associated with a heightened risk of severe disease or death, as have certain germline genetic variants. However, no one or combination of these factors fully explain the heterogeneity in outcomes observed with the disease.

A large number of biomarkers have been studied for their potential utility of predicting a more or less severe clinical course of COVID-19, including IL-6, IL-2R, IL-8, IL-10, CRP, PCT, and TNF alpha [1], but so far none have proven sufficiently accurate to help in triaging or managing COVID-19 infected patients. In addition, a number of HLA alleles [2] and several variants of the *ACE2* [3] and *TMPRSS2* [4] genes affecting the expression of the receptors related to COVID-19 have been associated with the disease susceptibility; and two genome-wide association studies have identified loci associated with disease severity [5]. Overall, the magnitudes of effect reported in these studies are modest, considerable heterogeneity across studies was observed, and concerns about inappropriate use of this information have recently been raised [6]. In addition, several studies have investigated changes in the plasma proteome in conjunction with COVID-19 [7-9], demonstrating that a range of proteins, primarily those associated with neutrophil activation, complement activation, platelet function, and T cell suppression, as well as of proinflammatory factors upstream and downstream of interleukin-6, interleukin-1B, and tumor necrosis factor showed significant differential expression in severe compared to asymptomatic or mild disease.

Given the fact that protein glycosylation is commonly observed to undergo changes in a range of medical conditions, it was of interest to study the glycoproteome in the setting of COVID-19, and to compare potential profile differences as they may be found in individuals who had experienced a severe disease course rather than an asymptomatic one. Also, we were interested in contrasting these attributes with the glycoproteome profiles of other comparison groups, including individuals with an indolent coronavirus-related common cold illness, healthy controls with no evidence of coronavirus exposure, and individuals with bacterial sepsis. With regard to the latter, we argue that the contrast between two systemic inflammatory syndromes may shed additional light on COVID-19-specific phenomena. In sepsis, similar to COVID-19, pro-inflammatory and anti-inflammatory mediators such as tumor necrosis factor-α (TNF-α), interleukin-1β (IL-1β), interleukin-6 (IL-6), and monocyte chemoattractant protein 1 (MCP-1) [10] are released, followed by a rise in the levels of acute phase proteins such as procalcitonin, calprotectin, pro-adrenomedullin, pentraxin-3, and C-reactive protein (CRP) [11]. While a number of studies interrogating plasma glycoprotein isoforms in severe systemic inflammatory states have been published in the context of bacterial sepsis, describing among others, changes of alpha-1-antichymotypsin and IgG [12-17] glycosylation following a septic episode, distinguishing survivors from patients who died, or differentiating febrile individuals with and without bacteremia, no such studies have so far been published in the context of COVID-19.

We deployed to this task a recently developed glycoprotein profiling technology platform that couples high-resolution liquid chromatography (LC)-mass spectrometry (MS) with an artificial intelligence (AI), neural network (NN)-based high-throughput data processing software, which has allowed us to scale previously labor intensive glycoproteomic analysis for accurate quantification of site-specific protein glycosylation by several orders of magnitude. This platform has enabled us to identify predictive glycoproteomic signatures in a range of clinical conditions.

## 2. Materials and Methods

### 2.1. Biological Samples

The sample set consisted of 115 samples including 50 (39 serum, 11 plasma) from patients hospitalized with PCR-confirmed severe symptomatic COVID-19, 22 serum samples from individuals without a history of symptomatic COVID-19 illness who were found to be seropositive for SARS-CoV-2-antibodies when they presented as blood bank donors (here called “asymptomatic COVID-19”), 16 plasma samples from patients who had presented with bacterial sepsis (8 mild, 8 severe), 12 plasma samples from patients who were positive by PCR for a common cold-presenting coronavirus, and 15 serum samples of healthy, coronavirus seronegative controls. Samples of the latter three groups had been collected well before the COVID-19 pandemic. All samples were provided fully deidentified, with samples from severely ill COVID-19 patients being remnants of specimens collected for routine clinical care or analysis (left-over specimens), thus demographic data were not available in a subset of these samples. For the 62 of 72 COVID-19 patients for which age and sex were known, symptomatic COVID-19 patients were on average 10 years older (mean = 58.5 years, SD = 13.0) than asymptomatic COVID-19 subjects (mean = 48.2 years, SD = 15.4). Among symptomatic COVID-19 patients with known sex, a majority – 25 of 40 – were male. Sepsis patients were, on average, 66.5 years old (SD = 14.3), and 11 of the 16 were male. Age and sex information was not known for the healthy control and common cold coronavirus samples; likewise, no information on preexisting medical conditions was known for any of the subjects included in the study (Table 1).

**Table 1.** Cohort summary, to the extent annotations were available.

| Phenotype | Source | N* | Serum | Plasma | Train set | Test set | Male | Female | Med. Age (IQR) |
| --- | --- | --- | --- | --- | --- | --- | --- | --- | --- |
| Symptomatic COVID-19+ | Kaiser Permanente | 50 | 39 | 11 | 38 | 12 | 25 | 15** | 55.5 (50.5, 67.3) |
| Bacterial sepsis | U of Florida, Jacksonville | 16 | 0 | 16 | 12 | 4 | 11 | 5 | 60.5 (57, 73.3) |
| Common cold coronavirus | Stanford Blood Bank | 12 | 0 | 12 | 9 | 3 | n/a | n/a | n/a |
| Asymptomatic COVID-19+ | Stanford Blood Bank | 22 | 22 | 0 | 16 | 6 | 10 | 12 | 49 (40.3, 61) |
| Healthy controls | Stanford Blood Bank | 15 | 15 | 0 | 11 | 4 | n/a | n/a | n/a |

\*Excluding 5 outliers; \*\*10 symptomatic COVID-19 patients do not have reported sex

### 2.2. Chemicals and reagents

Pooled human serum (for assay normalization and calibration purposes), dithiothreitol (DTT), and iodoacetamide (IAA) were purchased from MilliporeSigma (St. Louis, MO). Sequencing grade trypsin was purchased from Promega (Madison, WI). Acetonitrile (LC-MS grade) was purchased from Honeywell (Muskegon, MI). All other reagents used were procured from MilliporeSigma, VWR, and Fisher Scientific.

### 2.3. Preanalytical sample preparation

Serum samples were reduced with DTT and alkylated with IAA followed by digestion with trypsin in a water bath at 37°C for 18 hours. To quench the digestion, formic acid was added to each sample after incubation to a final concentration of 1% (v/v).

### 2.4. Liquid chromatography/mass spectrometry (LC-MS) analysis

Digested serum samples were injected into an Agilent 6495C triple quadrupole mass spectrometer equipped with an Agilent 1290 Infinity ultra-high-pressure (UHP)-LC system and an Agilent ZORBAX Eclipse Plus C18 column (2.1 mm internal diameter x 150 mm length, 1.8 µm particle size). Separation of the peptides and glycopeptides was performed using a 70-min binary gradient. The aqueous mobile phase A was 3% acetonitrile, 0.1% formic acid in water (v/v), and the organic mobile phase B was 90% acetonitrile 0.1% formic acid in water (v/v). The flow rate was set at 0.5 mL/min. Electrospray ionization was used as the ionization source and was operated in positive ion mode. The triple quadrupole MS was operated in dynamic multiple reaction monitoring (dMRM) mode. Samples were injected in a randomized fashion with regard to underlying phenotype, and reference pooled serum digests were injected interspersed with study samples to allow for correction of within-run drift of baseline signal. For quality control purposes, the ratios of glycopeptide abundance relative to their cognate non-glycosylated peptides were assessed in pooled serum replicates by run order. Five representative system suitability glycopeptide biomarkers from each abundance category were monitored, for a total of 15; 10 of these 15 glycopeptides had a coefficient of variation below 10%, while 14 were below 20%.

### 2.5. Data analysis

We quantified 728 peptides and glycopeptide isoforms, reflected by 1013 MRM transitions, and representing 73 high-abundance (concentrations of >10 µg/ml) serum glycoproteins. Our transition list consisted of glycopeptides as well as of non-glycosylated peptides from each glycoprotein. To build machine learning models, the R libraries ‘stats’ and ‘caret’ [18] were used. We used PB-NET [19], a peak integration software that had been developed in-house, to integrate peaks and obtain raw abundances for each biomarker (i.e., peptides and glycopeptides).

Normalized abundances of glycopeptides and peptides among groups of patients with severe and asymptomatic COVID-19, sepsis, common cold coronavirus, and healthy controls were assessed. The raw abundance features for each biomarker were normalized by using spiked-in heavy-isotope-labeled internal standards with known peptide concentrations, where available (see below). The calculation relies either on relative abundance when only one or two glycans are present at a site, i.e. the quotient of raw abundance signal intensity of the glycopeptide(s) and the raw abundance of a corresponding non-glycosylated peptide from the same protein, or on site occupancy when more than two glycan moieties are present at a given glycosylation site, i.e. on the fractional abundance across all glycans observed at that site. For each glycopeptide biomarker, the product of its site occupancy or relative abundance and corresponding peptide concentration is used to calculate approximate glycopeptide concentration. This is what is described as normalized abundance at later points in this paper. If an internal standard was not available for a particular protein, a surrogate was used instead, chosen by the similarity of the protein’s m/z value to one of the available internal standards. Concentration data for 531 glycopeptides, 320 of which are based on site occupancy and 211 on relative abundance, and for 66 peptide biomarkers were ultimately used for the analysis, totaling 597 unique biomarkers.

An additional correction factor was included to account for the differences in signal intensity encountered between plasma and serum samples. Using the serum samples from symptomatic COVID-19 patients as a reference and comparing them to plasma samples from the same group of patients, marker-specific multiplicative factors were derived and applied to all plasma samples, resulting in a reduction of the distance encountered among clusters for plasma and serum samples that were observed in the uncorrected principal component analysis. A visualization of the principal component analysis factoring in the plasma-serum correction factor is shown in Figure 1.

**Figure 1.**
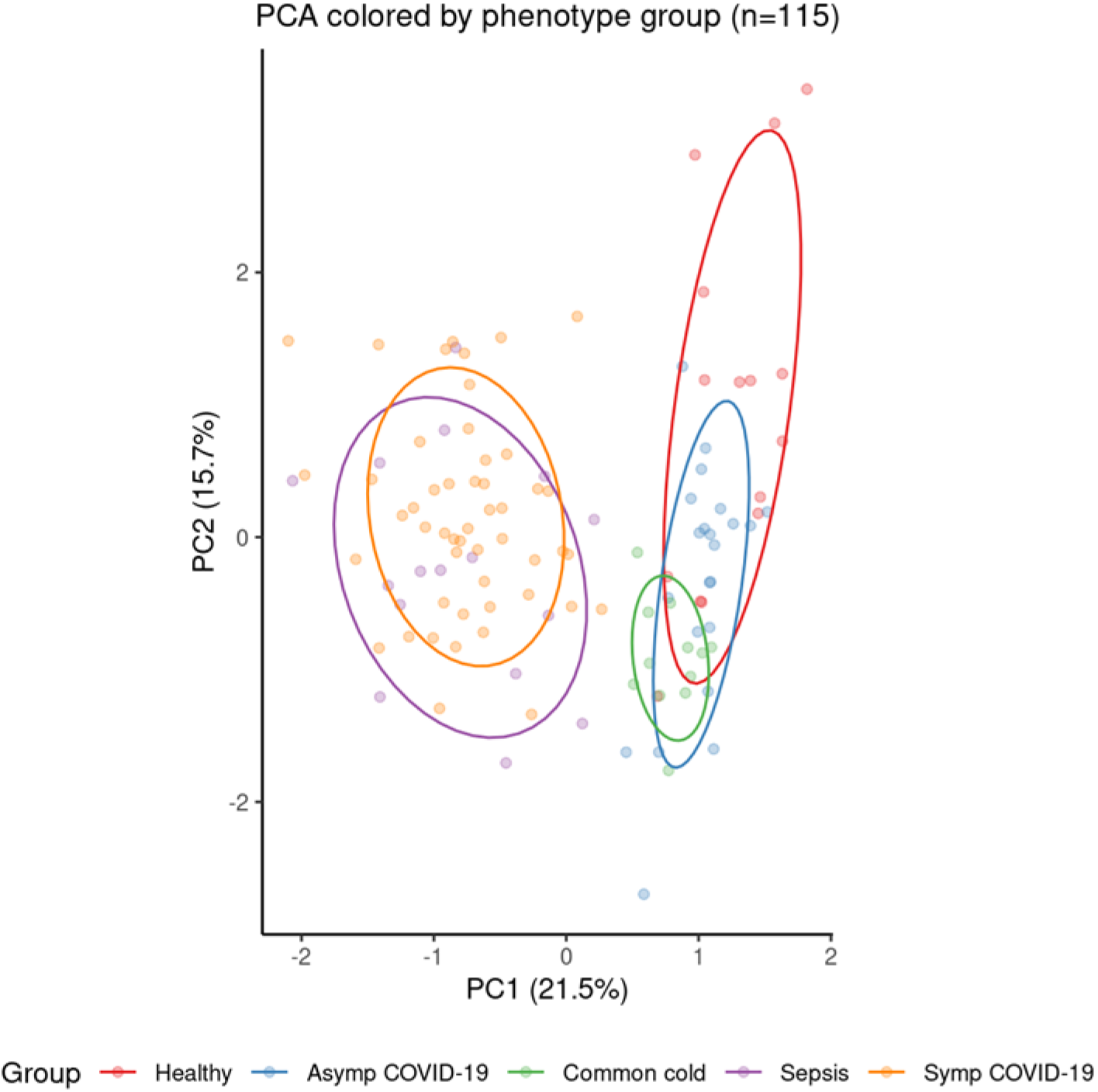
Visualization of top two principal components in PCA of all 115 subjects included in the analysis (subjects are colored by phenotype).

To compare any two phenotype groups, we used linear regression on a feature-by-feature basis with phenotype serving as the sole binary independent variable. Analyses were not adjusted for age and sex for the sake of consistency since the respective data were not available for samples of all phenotype groups. Of note, when age and sex adjustments were made for those phenotype groups in which data were available, results were not statistically significantly different compared to the same analyses with these terms removed (results not shown). Correcting for multiple comparisons (i.e., number of biomarkers analyzed simultaneously), differences of any biomarker among phenotype groups compared were considered statistically significant if they satisfied a false discovery rate (FDR) of less than 0.05. Overlapping sets of significant biomarkers between sets of groups were then assessed.

For supervised multivariate modeling, features were log-transformed and split into a training (n=86) and a test set (n=29). To perform binary classification, repeated five-fold cross-validated LASSO-regularized logistic regression was used to predict probability of symptomatic COVID-19 with hyperparameters tuned to prevent overfitting and promote balanced sensitivity and specificity metrics. For unsupervised multi-class classification, K-means clustering methods were performed on all 115 patients without using the training or testing sets to predict group membership in three distinct groups - symptomatic COVID-19, sepsis, or other.

### 2.6. Pathway Analysis

To identify the most relevant canonical pathways related to the findings of our study, we used Ingenuity® Pathway Analysis software (QIAGEN Inc.) among healthy control subjects, asymptomatic COVID-19 cases, and symptomatic COVID-19 patients. We analyzed glycoproteins which demonstrated statistically significant differences in either protein abundance or glycosylation levels between the symptomatic COVID-19 and healthy control groups. Deregulated canonical pathways were identified at p-value < 0.01. Upstream regulators of 23 acute phase glycoproteins were predicted with a molecular type filter including only genes, RNAs, and proteins. The protein networks associated with these differentially abundant glycoprotein biomarkers were automatically generated with both direct and indirect relationships.

## 3. Results

### 3.1. Logistic regression results comparing individual phenotype groups

Logistic regression analysis revealed a large number of statistically significantly different normalized biomarker abundances between individual phenotype groups. Many of these overlap between multiple contrasts when using healthy controls as the reference phenotype group; these are summarized in Figure 2. Likewise, volcano plots are presented in Figure 3, and an overall heatmap in Figure 4.

**Figure 2.**
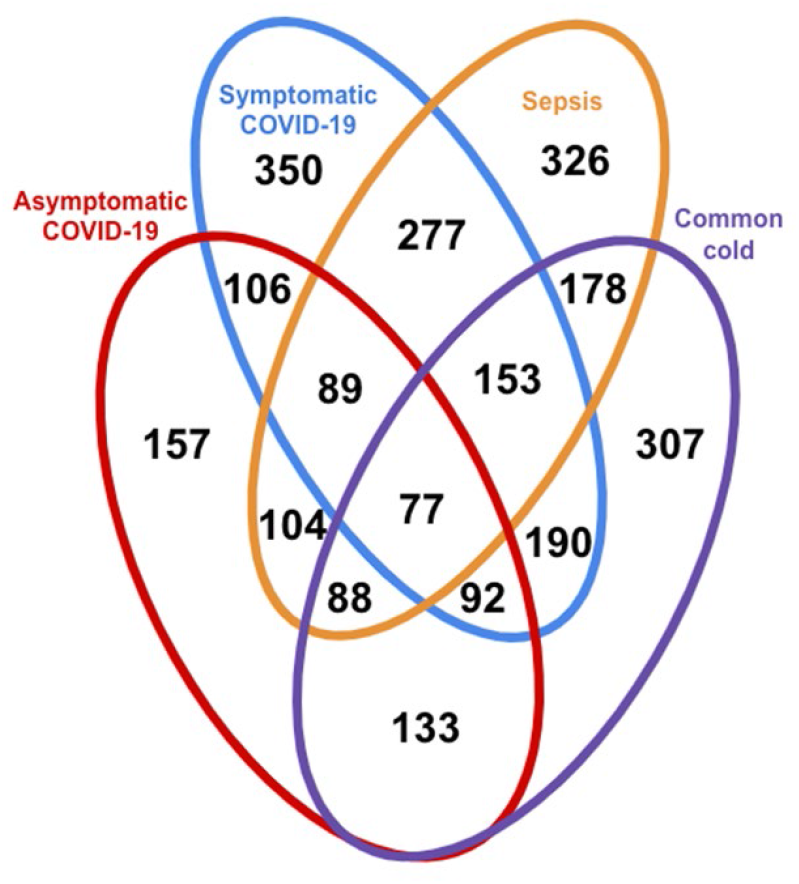
Venn diagram indicating number of differentially expressed biomarkers at FDR<0.05 between healthy controls and given phenotype group(s).

**Figure 3.**
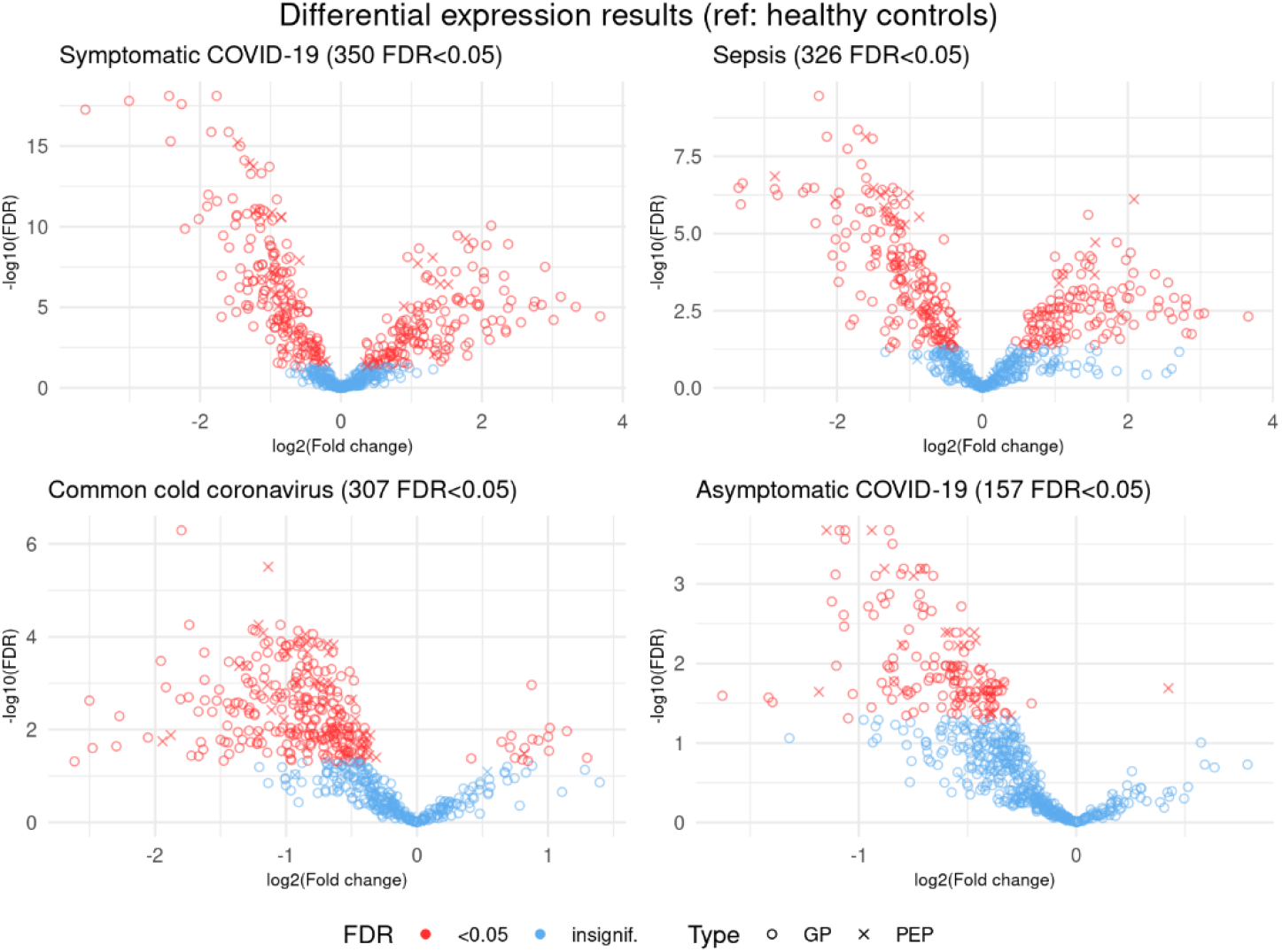
Volcano plots showing log-transformed multiplicative fold changes and respective log-transformed false discovery rates (FDR) for each biomarker in differential expression analysis with healthy controls used as the reference for each phenotype. Biomarkers in red represent those that are statistically significantly differentially expressed at FDR<0.05. Biomarkers marked with a circle are glycopeptides, while an X represents non-glycosylated peptides.

**Figure 4.**
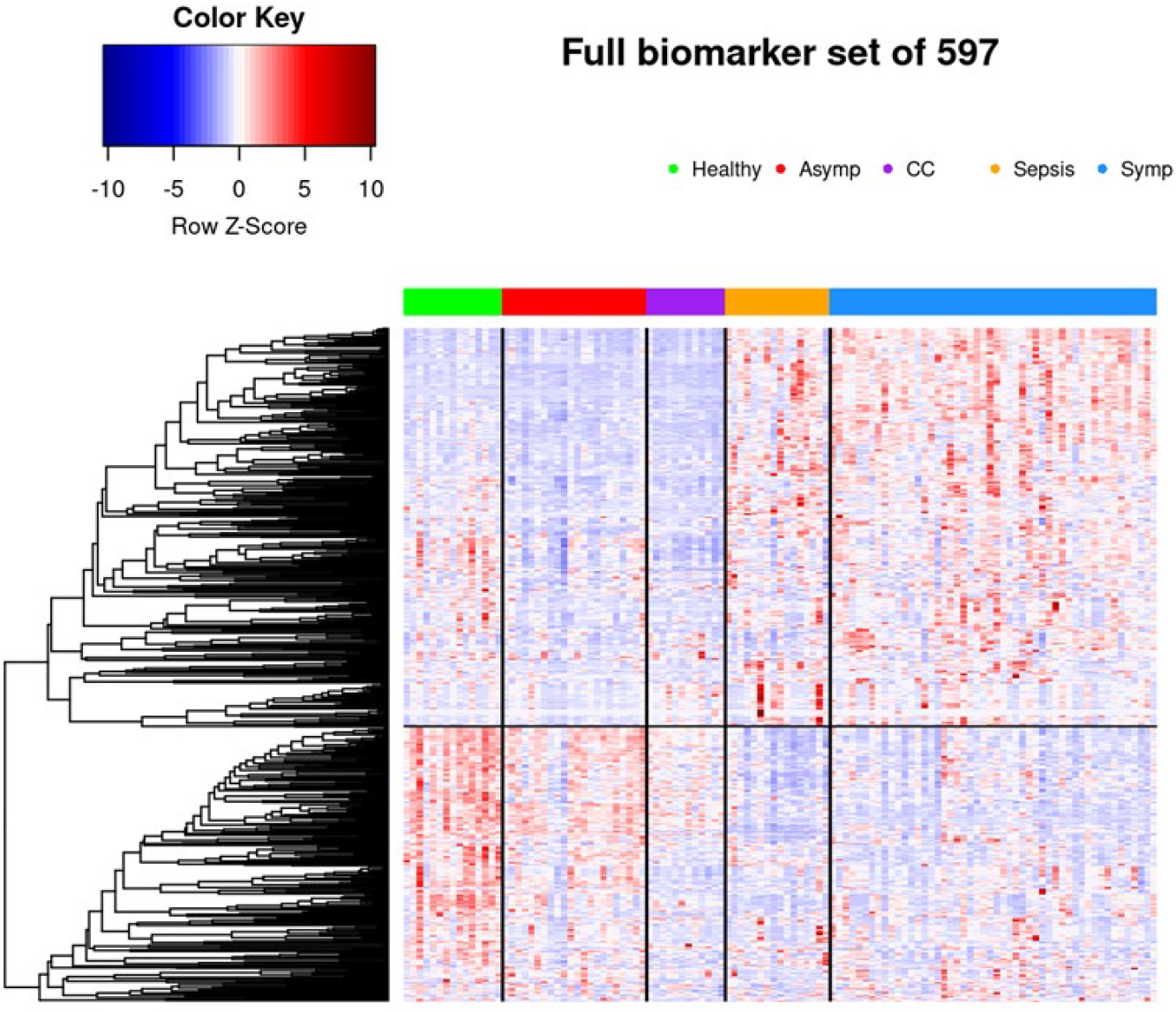
Heatmap in which all 115 patients and 597 biomarkers are represented, clustered into phenotype groups column-wise and hierarchically clustered row-wise, respectively. Row-wise Z-scores determine the color of each cell.

#### 3.1.1. Comparison of healthy control samples to other groups

Among the healthy controls and symptomatic COVID-19 patients, 350 glycopeptides and peptides were statistically significantly differently expressed at FDR<0.05, as were 326 biomarkers among the healthy samples and those with sepsis, with 277 overlapping between the two phenotype contrasts. Among samples from healthy controls and individuals seropositive for common cold coronavirus, 307 biomarkers were statistically significantly differentially expressed, 153 of which were also differentially expressed among healthy controls and both symptomatic COVID-19 samples and sepsis samples. A comparatively smaller set of 157 biomarkers differed statistically significantly among healthy subjects and asymptomatic COVID-19 subjects (Figure 2), of which 77 also showed statistically significant differences among healthy controls and each of the other three groups (Figure 5).

**Figure 5.**
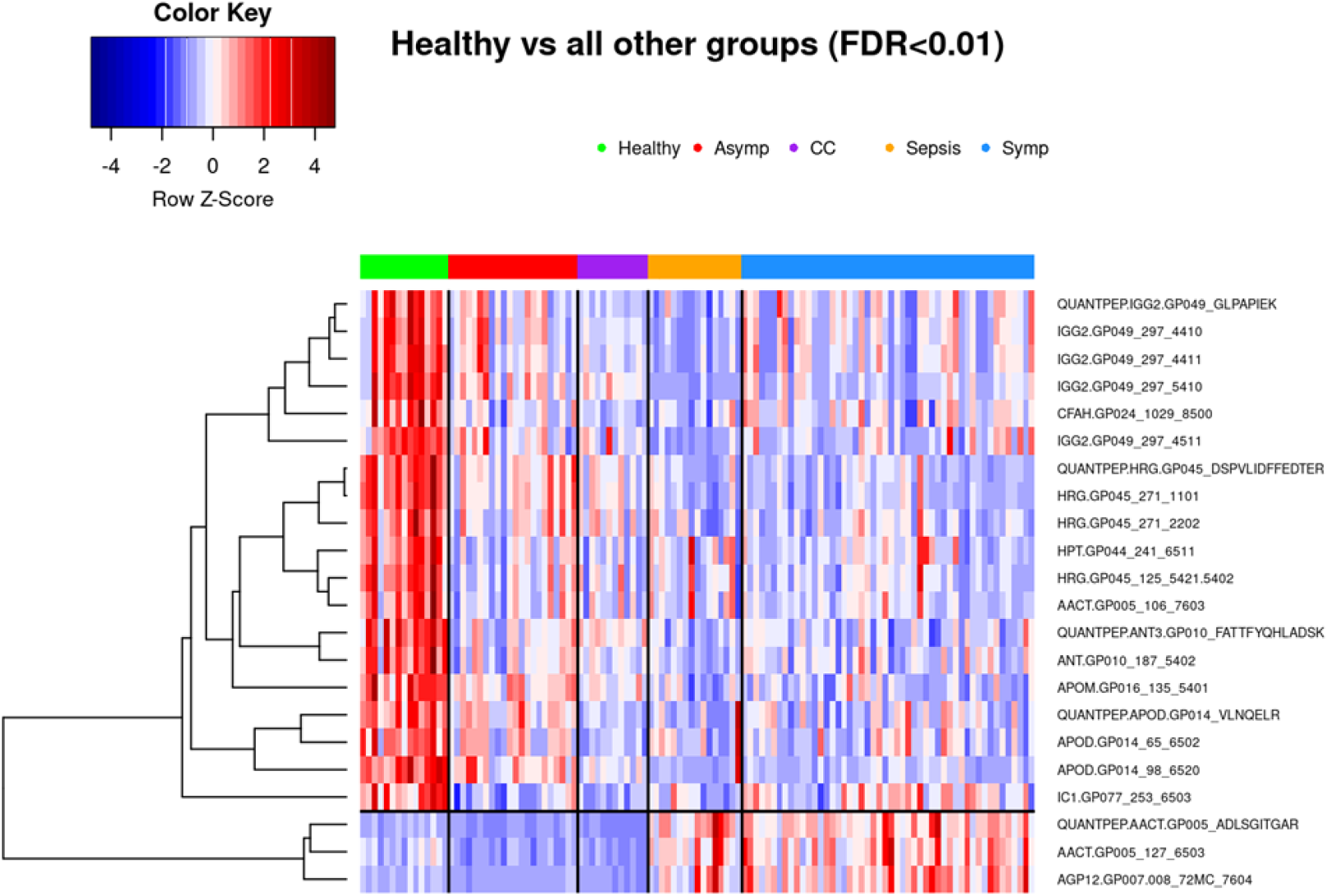
22 biomarkers that achieve FDR<0.01 in differential expression analysis between healthy controls and all four of the other phenotype groups separately. 77 biomarkers achieve FDR<0.05; a more conservative threshold was chosen for clarity of the heatmap. Subjects are clustered into phenotype groups column-wise and hierarchically clustered row-wise, respectively. Row-wise Z-scores determine the color of each cell.

#### 3.1.2. Comparison of symptomatic and asymptomatic COVID-19 samples

Among samples from symptomatic and asymptomatic COVID-19 subjects, 374 features showed statistically significant differences, with a substantial overlap of 272 markers among the 350 that were statistically significantly different when comparing samples of symptomatic COVID-19 and healthy controls (Figure 2).

#### 3.1.3. Comparison of symptomatic COVID-19 and sepsis samples

Glycoprotein abundance profiles showed striking similarities among samples of symptomatic COVID-19 and sepsis patients as illustrated in the respective principal component analysis (Figure 1) and heatmap (Figure 4). Of the 277 biomarkers that were statistically significantly differentially expressed between healthy controls and both symptomatic COVID-19 and sepsis, 276 were directionally concordant. Of the 114 biomarkers that were statistically significantly upregulated in symptomatic COVID-19 and sepsis compared to healthy controls, 63 (55.3%) have a more extreme fold change in symptomatic COVID-19 patients, whereas among the 162 biomarkers statistically significantly down-regulated in both symptomatic COVID-19 and sepsis, 128 (79.0%) have a more extreme fold change in sepsis patients. Of note, we found 101 biomarkers – 65 with higher, and 36 with lower abundance in COVID-19 as compared to sepsis patients – to be statistically significantly different among samples from COVID-19 and sepsis patients, pointing to attributes distinguishing the two phenotypes (Figure 6 shows most significant subset). Additionally, 34 features were statistically significantly different among symptomatic COVID-19 samples as compared to any of the other phenotypes, thus representing a signature unique to this phenotype (Figure 7). Another set of 46 features were statistically significantly different among sepsis samples as compared to any of the other phenotype groups, representing a sepsis-specific signature (Figure 8). Among the 66 non-glycosylated peptides (all representing different proteins) assayed, 25 were statistically significantly downregulated in both symptomatic COVID-19 and sepsis at FDR<0.05, while only seven were statistically significantly upregulated in both (Figure 9).

**Figure 6.**
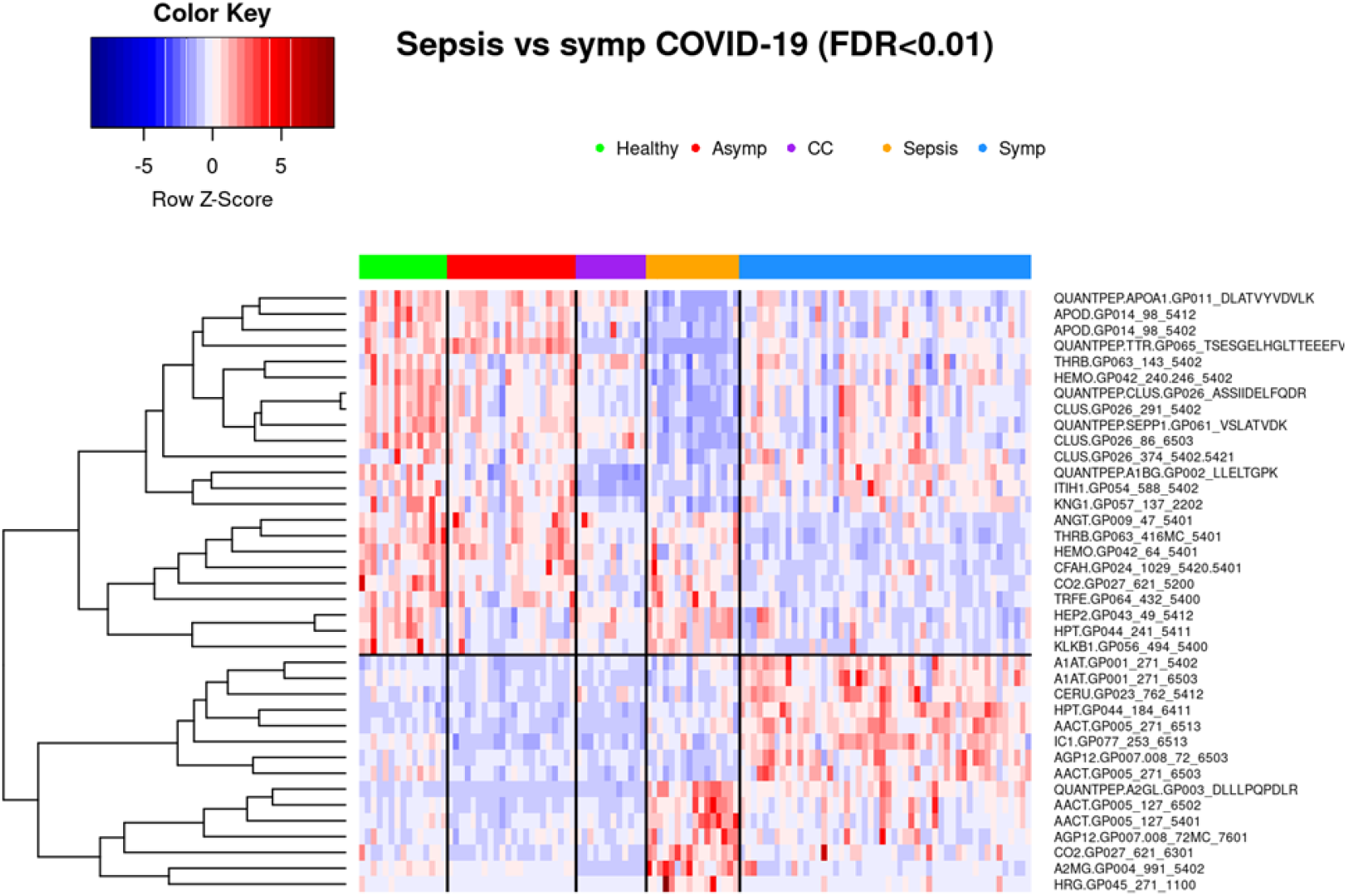
38 biomarkers that achieve FDR<0.01 in differential expression analysis between bacterial sepsis and symptomatic COVID-19 patients. 101 biomarkers achieve FDR<0.05; a more conservative threshold was chosen for clarity of the heatmap. Subjects are clustered into phenotype groups column-wise and hierarchically clustered row-wise, respectively. Row-wise Z-scores determine the color of each cell. CC: Common cold coronavirus

**Figure 7.**
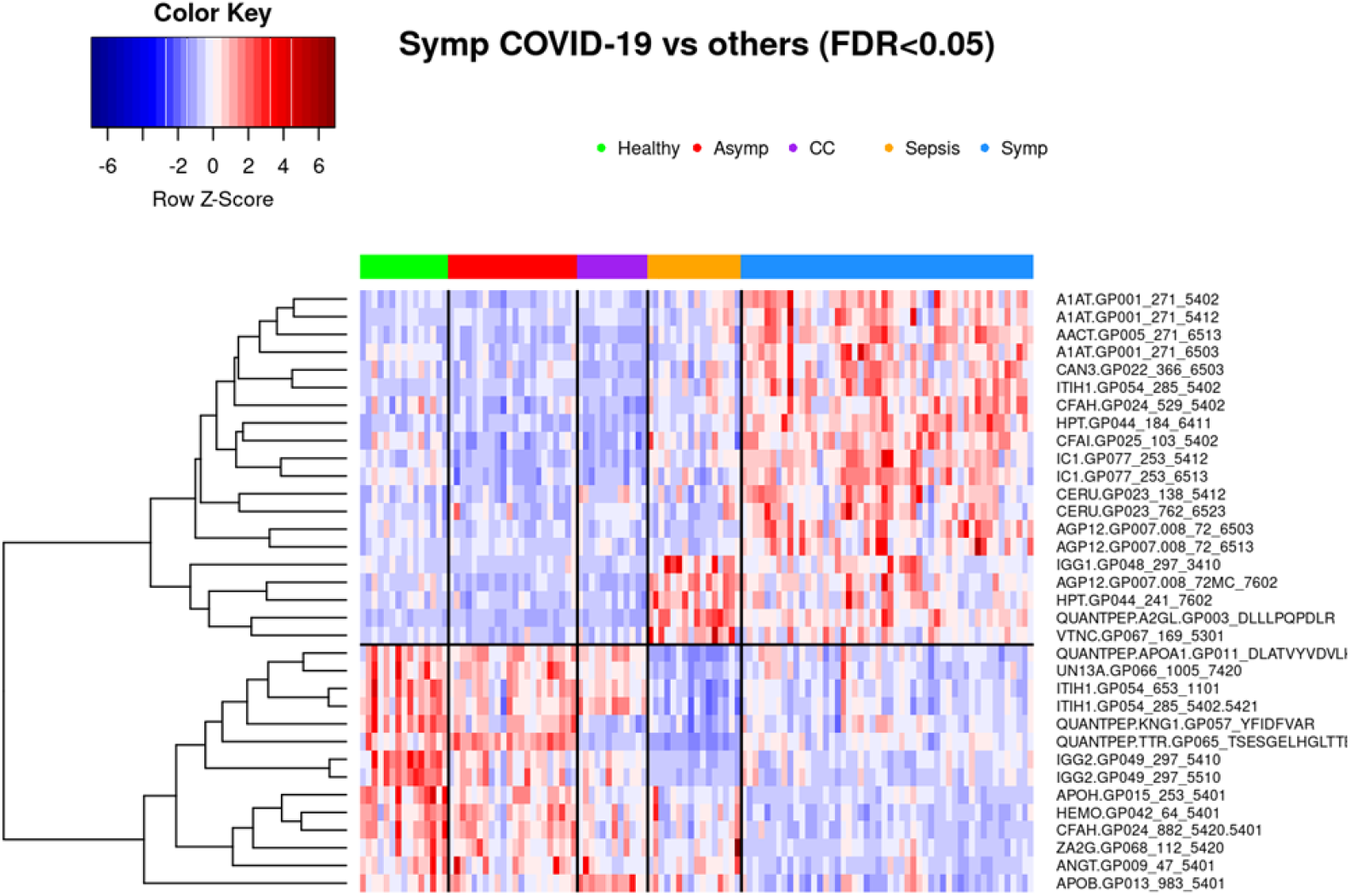
34 biomarkers that achieve FDR<0.05 in differential expression analysis between symptomatic COVID-19 and all four of the other phenotype groups separately. Subjects are clustered into phenotype groups column-wise and hierarchically clustered row-wise, respectively. Row-wise Z-scores determine the color of each cell. CC: Common cold coronavirus

**Figure 8.**
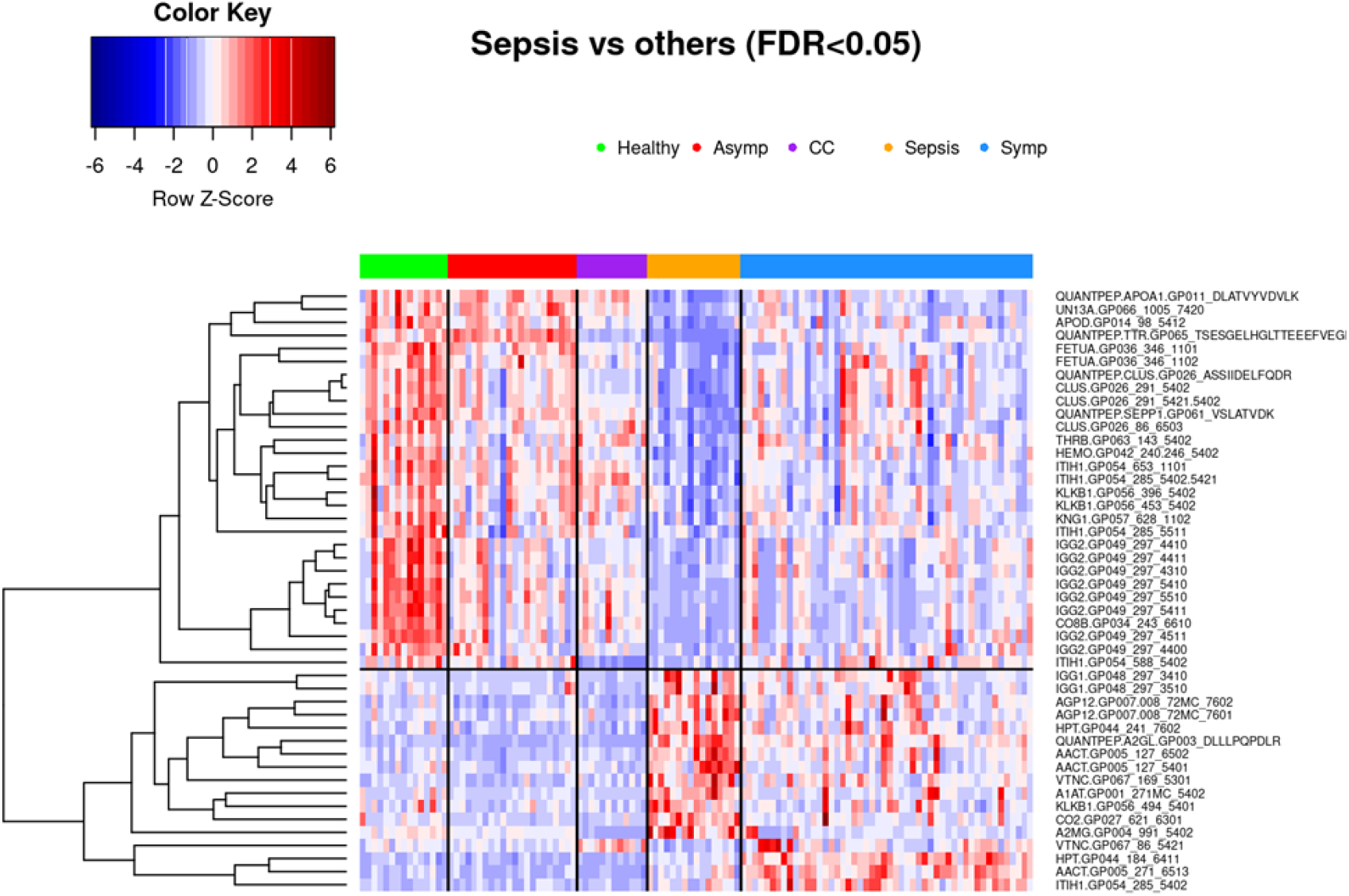
46 biomarkers that achieve FDR<0.05 in differential expression analysis between sepsis and all four of the other phenotype groups separately. Subjects are clustered into phenotype groups column-wise and hierarchically clustered row-wise, respectively. Row-wise Z-scores determine the color of each cell. CC: Common cold coronavirus

**Figure 9.**
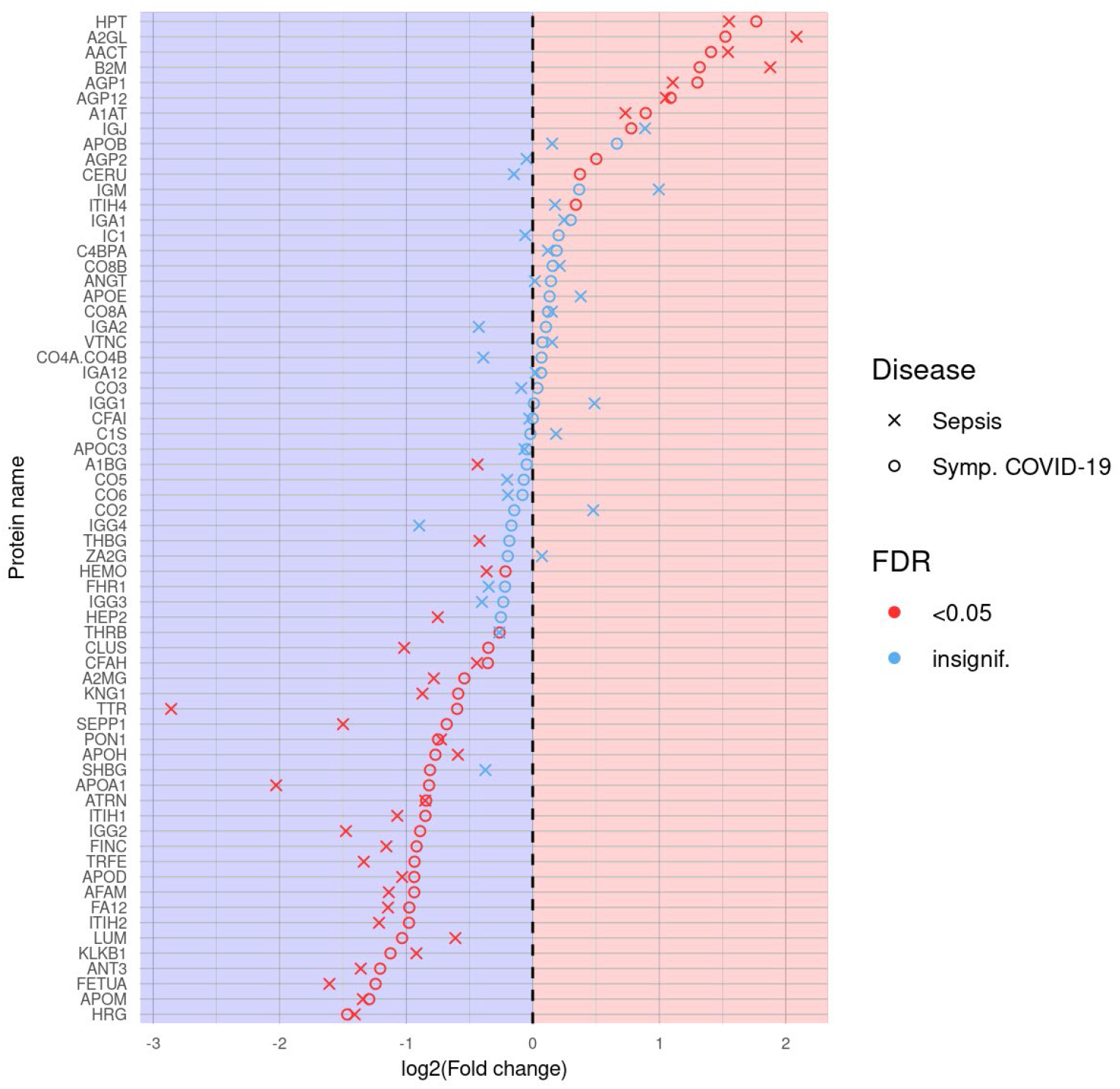
Dotplot showing log-transformed fold changes for each non-glycosylated peptide, using healthy controls as the reference against the sepsis and symptomatic COVID-19 (by which this is sorted) phenotype groups, each indicated using its own symbol. Red symbols represent those that are statistically significant at FDR<0.05.

### 3.2. Glycoproteomic signatures predicting symptomatic COVID-19 and other phenotype status

#### 3.2.1. Classification of symptomatic COVID-19, sepsis, and other samples using K-means clustering

As indicated by a plot showing the first two principal components according to group membership (Figure 1), and by a heatmap of all patients clustered by group and all standardized features (Figure 4), it is evident that patients with symptomatic COVID-19 and sepsis have a drastically different glycoproteomic signature as compared to those with asymptomatic COVID-19 or common cold coronavirus exposure, as well as healthy controls. When clustering the three control groups into one large group and removing their labels, the unsupervised K-means clustering algorithm provides 96% classification accuracy in allocating all 115 patients to one of three distinct groups based on the set of 34 features that statistically significantly differentiate symptomatic COVID-19 patients from all other phenotype groups at FDR<0.05 in the full dataset (Figure 7). Despite the absence of a training set, the full data naturally separates into these three clusters with a high degree of accuracy: 100% of the group comprising asymptomatic COVID-19, common cold coronavirus, and healthy control samples are allocated to cluster 1, 88% of sepsis patients are allocated to cluster 2, and 94% of symptomatic COVID-19 patients are allocated to cluster 3 (Figure 10, Table 2). The only five misclassifications observed were sepsis and symptomatic COVID-19 patients being mistaken for one another, respectively.

**Table 2.**
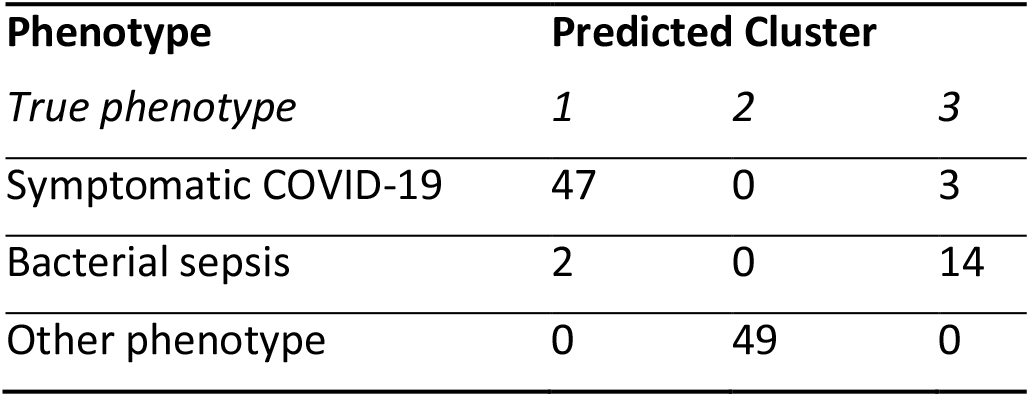
Allocation to predicted clusters based on K-means clustering

| Phenotype | Predicted Cluster |  |  |
| --- | --- | --- | --- |
|  | 1 | 2 | 3 |
| *True phenotype* |  |  |  |
| Symptomatic COVID-19 | 47 | 0 | 3 |
| Bacterial sepsis | 2 | 0 | 14 |
| Other phenotype | 0 | 49 | 0 |

**Figure 10.**
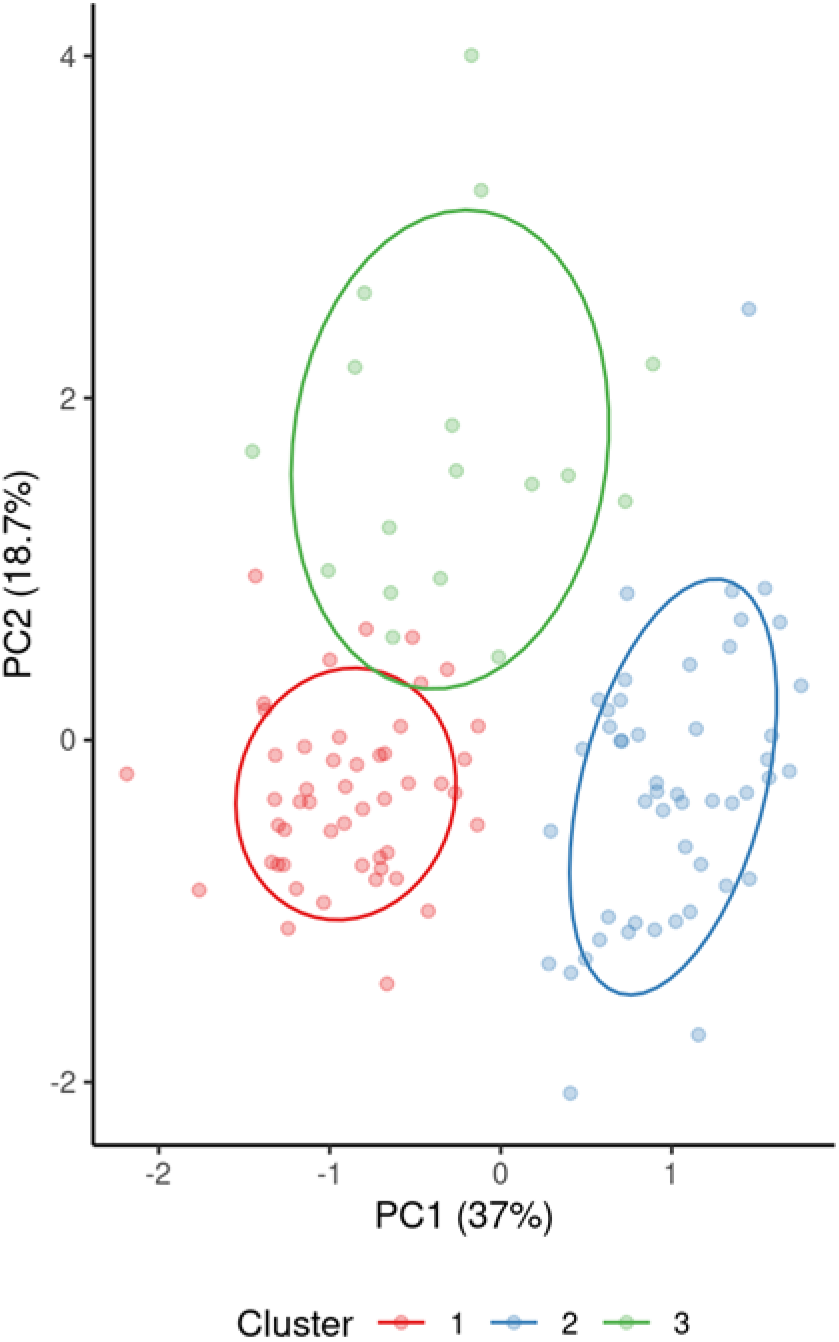
Results from K-means clustering, using only the 34 biomarkers that statistically significantly differentiate symptomatic COVID-19 patients from all of the other phenotype groups, visualized via principal component analysis.

#### 3.2.2 Classification of symptomatic COVID-19 using LASSO regression

When predicting whether a sample belongs to a symptomatic COVID-19 patient or not, repeated five-fold cross-validated LASSO-regularized logistic regression was performed in a randomly selected training set stratified by phenotype. All 597 biomarkers were considered for coefficient shrinkage; the final model, which includes 16 glycopeptides and two non-glycosylated peptides, yields 100% accuracy (100% sensitivity and specificity) in both the training and test sets (Figures 11, 12). It should be noted that particular care was taken to promote generalization to unseen samples – in other words, avoid over-fitting and overparameterizing the final model. We await the acquisition of additional samples to further assess and validate this model.

**Figure 11.**
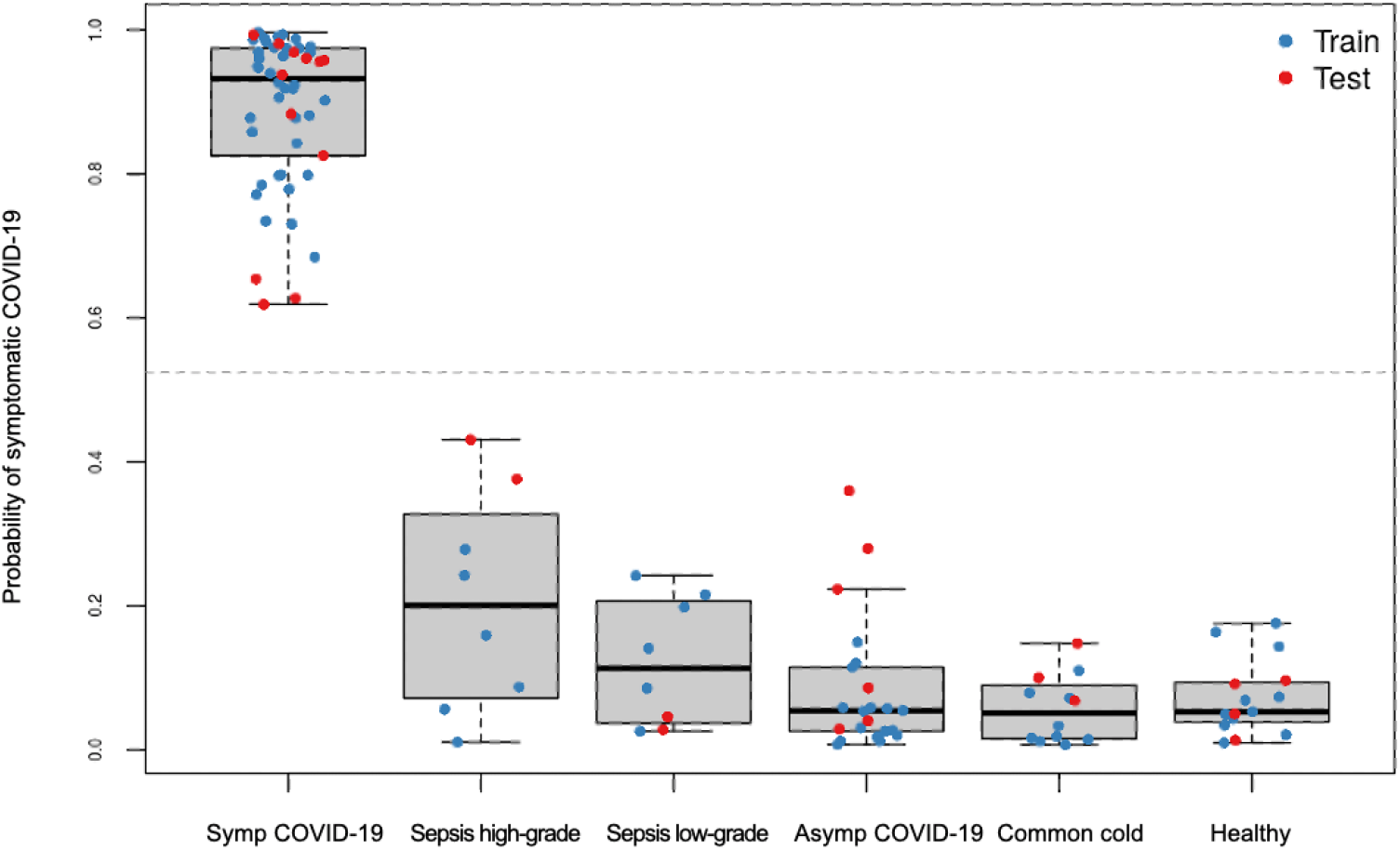
Predicted probabilities of symptomatic COVID-19 generated from LASSO-regularized logistic regression model, stratified by true phenotype group, and colored by training or testing set assignment.

**Figure 12.**
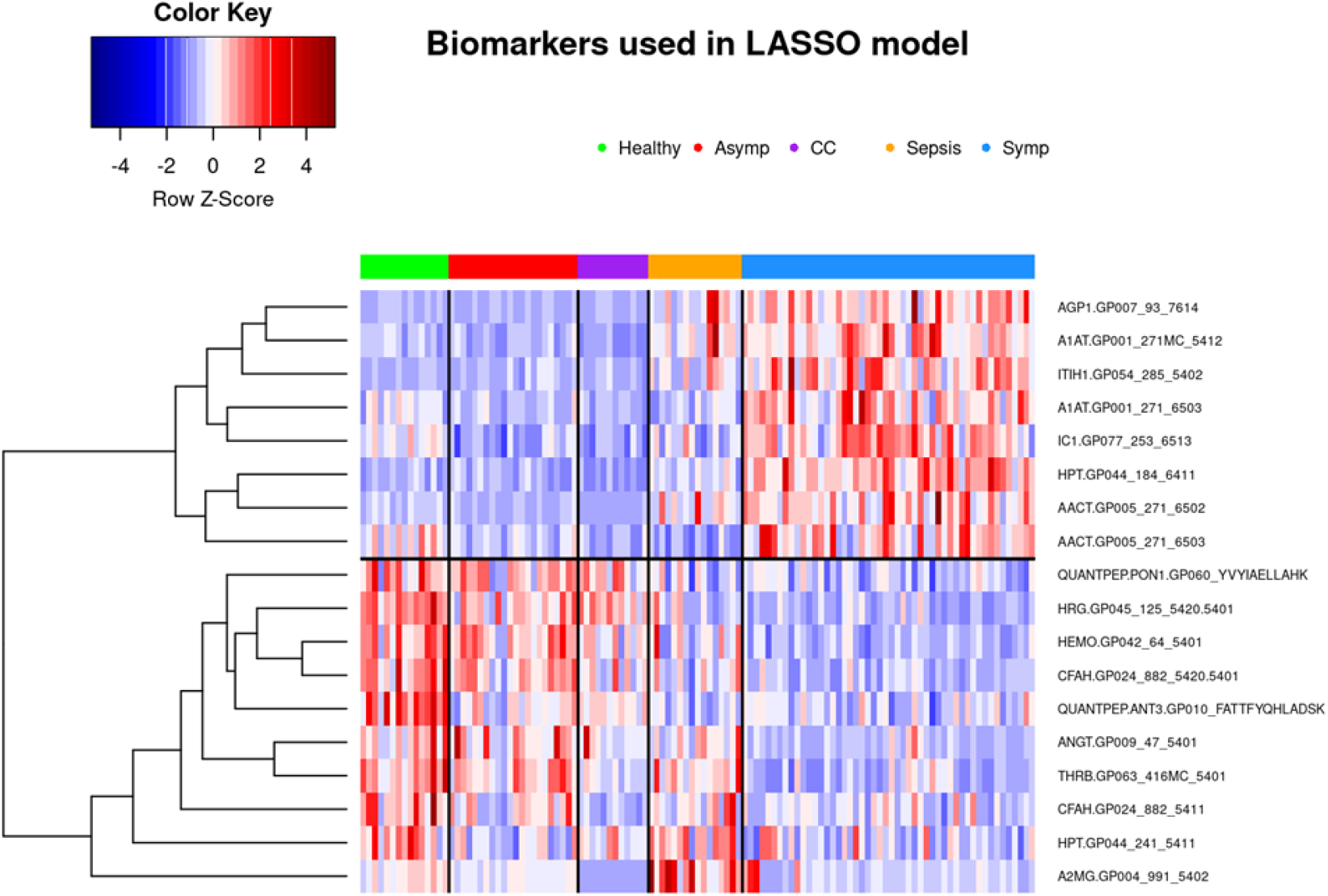
Heatmap showing retained biomarkers in LASSO-regularized classifier for all patients in both training and testing sets. Subjects are clustered into phenotype groups column-wise and hierarchically clustered row-wise, respectively. Row-wise Z-scores determine the color of each cell.

### 3.3. Bioinformatic analysis of observed findings

#### 3.3.1. Healthy vs. Symptomatic COVID-19

The 10 canonical pathways that were statistically most significantly (p<0.001) altered in symptomatic COVID-19 patients, compared with the healthy control group, are shown in Figure 13. Among these pathways, the acute phase response signaling pathway (p=1.1E-36) was identified as the most statistically significantly enriched pathway.

**Figure 13.**
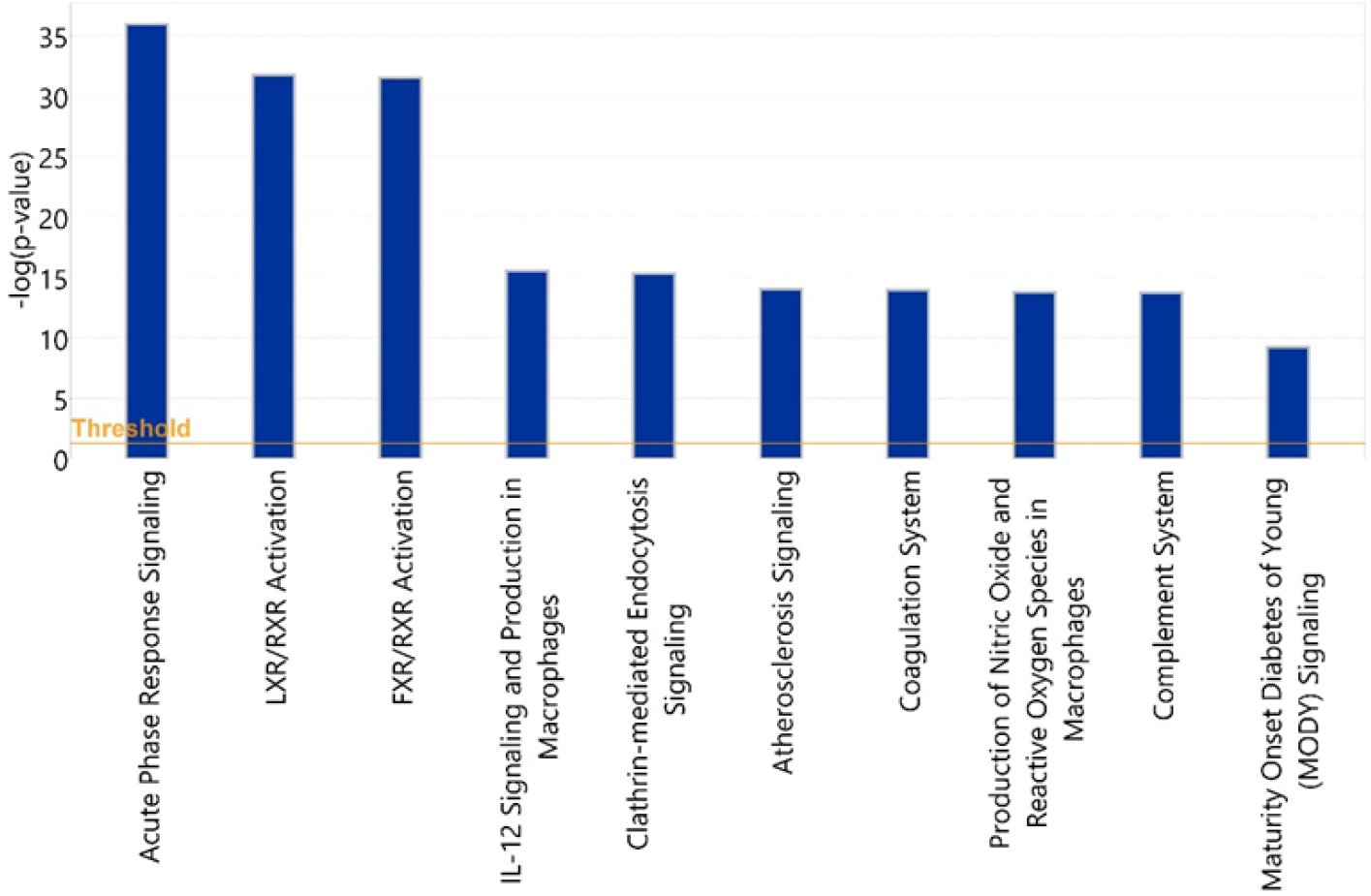
Statistical significance levels of differential activation of canonical pathways among healthy vs. symptomatic COVID-19 patients

Among the 58 input glycoproteins, 24 glycoproteins such as complement 3 (C3), alpha2 macroglobulin (A2M), haptoglobin (HP), hemopexin (HPX) and alpha-1-antichymotrypsin (SER-PINA3) are involved in this pathway. Two members of interleukin (IL)-6 cytokine family, IL-6 and Oncostatin M, were identified as most statistically significantly enriched upstream regulators of these identified acute phase proteins (Table 3 and Figure 14).

**Table 3.** Top 12 upstream regulators (Acute Phase Response Signaling)

| Upstream Regulator | Molecule Type | p-value of Target Molecules in Dataset | Target Molecules in Dataset |
| --- | --- | --- | --- |
| HNF1A | transcription regulator | 1.05E-14 | AGT, AHSG, APOH, ATP, C1S, C4BPA, F2, HPX, ITIH4, SERPINA1, SERPING1, TTR |
| IL6 | cytokine | 1.24E-13 | A2M, AGT, APOA1, ATP, CP, FN1, HP, HPX, ORM1, SERPINA1, SERPINA3, TF, TTR |
| HNF4A | transcription regulator | 1.1E-10 | AGT, AHSG, APOA1, APOH, ATP, C1S, CP, HPX, ITIH4, ORM1, ORM2, SERPINA1, SERPINA3, TF, TTR |
| Tcf 1/3/4 | group | 1.66E-08 | AHSG, APOH, TTR |
| Hmgn3 | other | 3.26E-08 | AHSG, APOA1, SERPINA1, TTR |
| OSM | cytokine | 4.43E-08 | A2M, C1S, C4BPA, FN1, HP, SERPINA1, SERPINA3, SERPING1 |
| CEBPB | transcription regulator | 8.56E-08 | AGT, CP, FN1, HP, HPX, ORM1, SERPINA1, TF |
| STAT1 | transcription regulator | 1.21E-07 | A2M, AGT, APOA1, C1S, FN1, SERPINA3, SERPING1 |
| STAT3 | transcription regulator | 1.66E-07 | A2M, AGT, AHSG, ATP, FN1, HP, SERPINA1, SERPINA3 |
| IL6ST | transmembrane receptor | 6.29E-07 | A2M, HP, HPX, ORM1 |
| TNF | cytokine | 1.13E-06 | A2M, AGT, APOA1, ATP, CP, FN1, HP, ORM1, SERPINA3, SERPIND1, TF |
| IL6R | transmembrane receptor | 1.58E-06 | A2M, FN1, HP, SERPINA3 |

**Figure 14:**
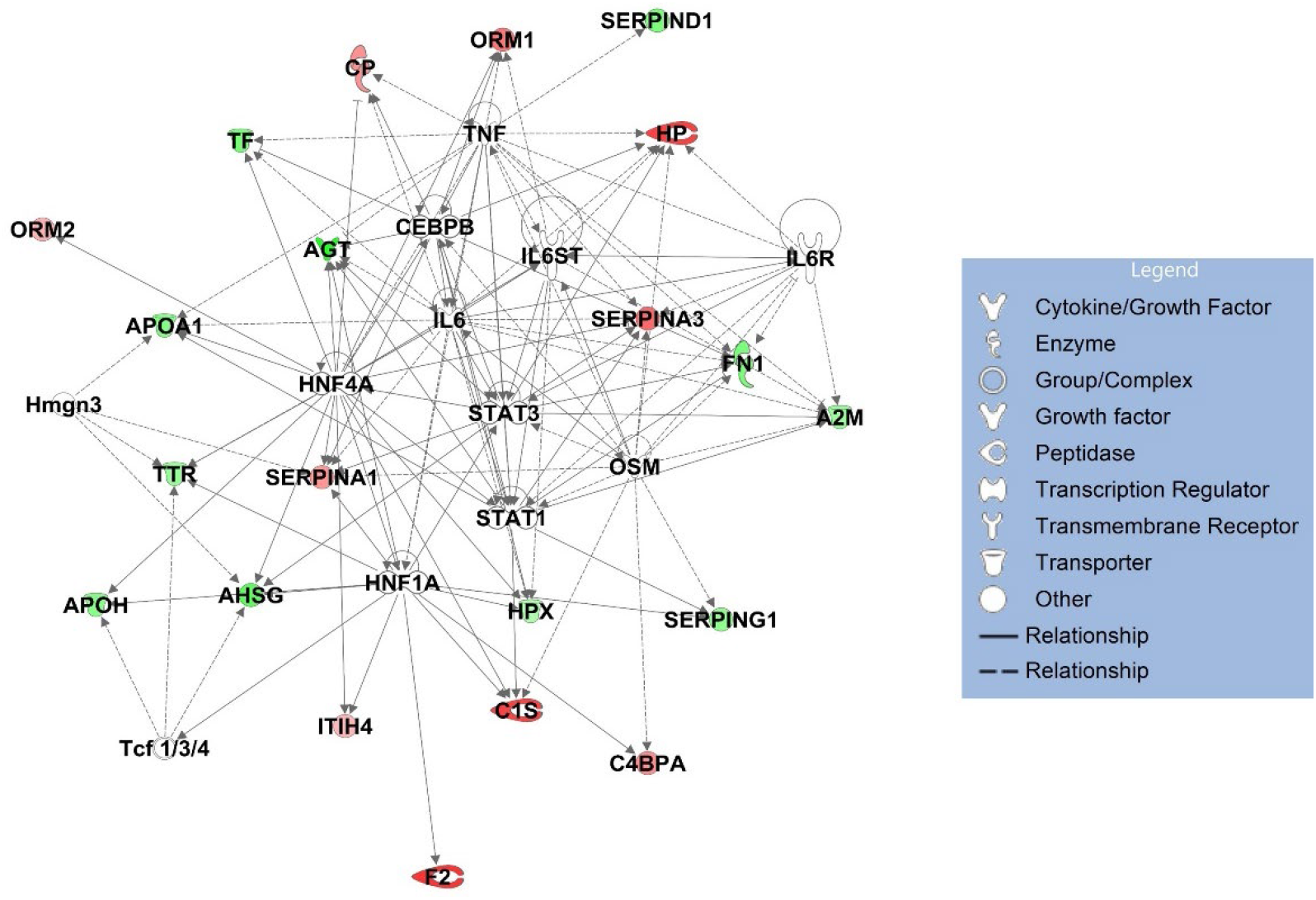
Network of 12 most statistically significantly altered upstream regulators (acute phase response signaling).

#### 3.3.2. Asymptomatic vs. Symptomatic COVID-19

To identify pathways associated with severity of COVID-19 illness, we applied IPA to analyze 64 glycoproteins which showed statistically significant differences in either protein abundance or glycosylation between symptomatic and asymptomatic COVID-19 patients. The 10 most statistically significant (p < 0.001) canonical pathways associated with disease severity are shown in Figure 15.

**Figure 15.**
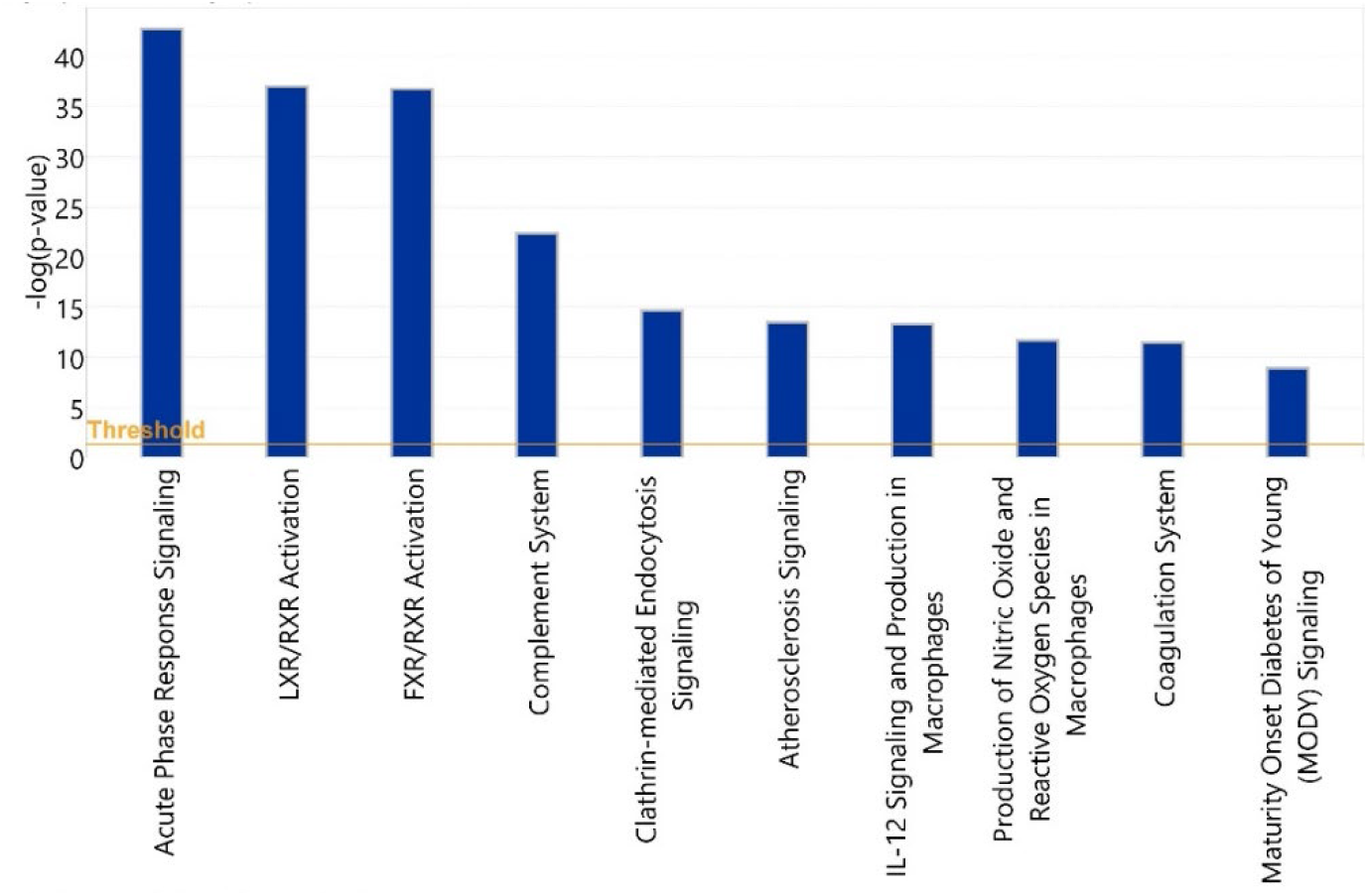
Statistical significance levels of differential activation of canonical pathways among asymptomatic vs. symptomatic COVID-19 patients.

These include acute phase response signaling, complement cascade, and the coagulation system pathways, representing 35 glycoproteins in total. Among the 35 glycoproteins, 28 glycoproteins are involved in the acute phase response signaling and 12 glycoproteins, including C3, C5, and C6, are involved in the complement system.

In our study, 28 acute phase glycoproteins were identified as statistically significantly differentially abundant in the symptomatic COVID-19 group compared with the asymptomatic COVID-19 group. Recently, Shen et al. [24] conducted a proteomic characterization of severe and non-severe COVID-19 patient sera. A comparison of our results and the Shen et al. findings shows that 11 acute phase proteins were detected by both studies (Figures 16 and 17). An additional 17 glycoproteins that are involved in the acute phase signaling pathway were identified by our study.

**Figure 16.**
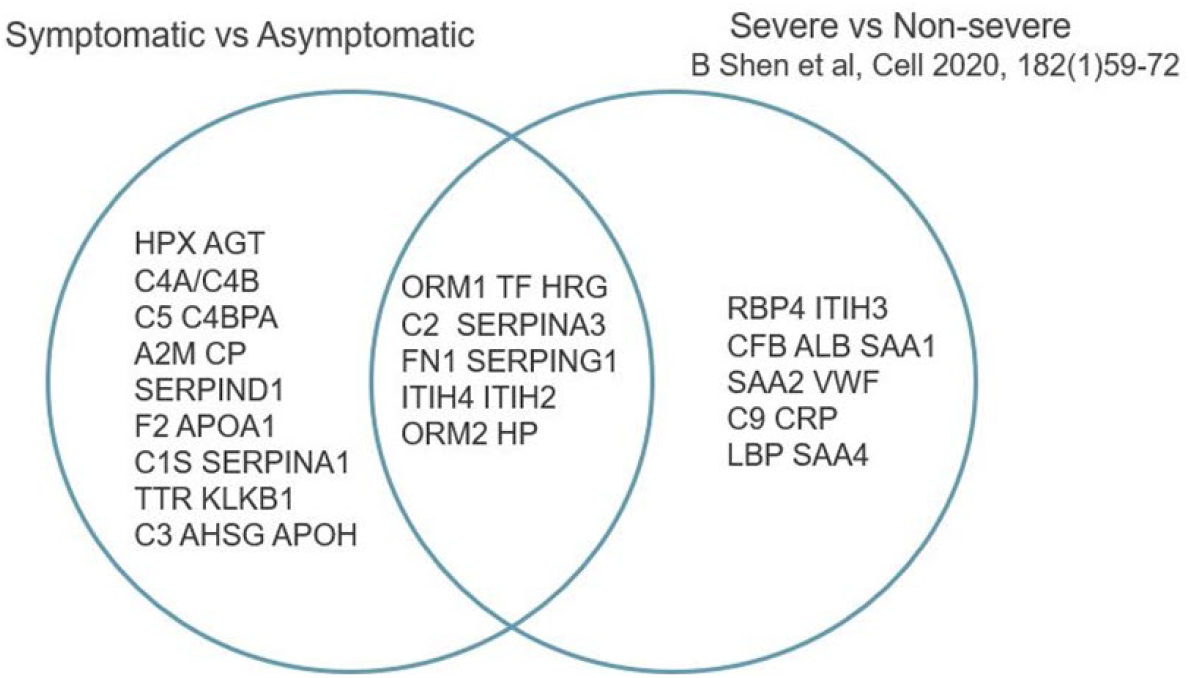
Acute phase proteins identified by this study and the study by Shen et al.

**Figure 17.**
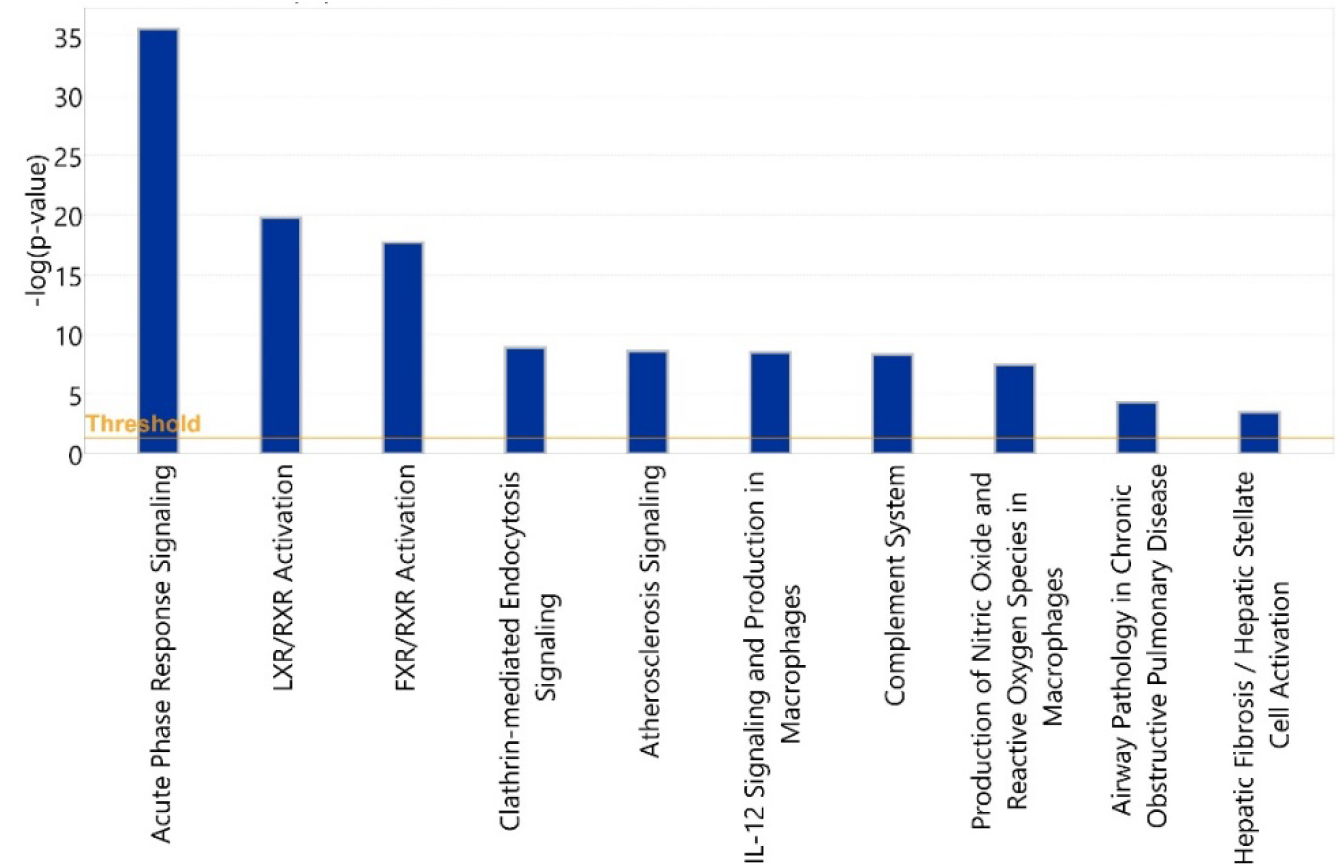
Statistical significance levels of differential activation of canonical pathways among non-severe vs. severe COVID-19 cases.

## 4. Discussion

In our study, samples from patients with symptomatic COVID-19 demonstrated glycoprotein profiles that are clearly different from those found in individuals who had experienced an asymptomatic or comparatively mild course of the disease. Comparing symptomatic COVID-19 with sepsis revealed a large number of corresponding changes, presumably indicative of non-specific changes associated with a severe systemic inflammatory state. However, we also observed a set of glycopeptides that displayed clear differences. While the concomitant changes observed in the two phenotypes are thus likely indicative of a secondary response to the inflammatory state resulting from either bacterial sepsis or COVID-19 infection, it is interesting to speculate whether the highly statistically significant predictive subset of 34 glycoforms that differentiate symptomatic COVID-19 and sepsis patients as well as the other control samples may represent a set of responses elicited specifically in patients suffering from a severe course of COVID-19, or possibly represent predisposing attributes associated with such a course of COVID-19 in contrast to a milder one.

The observation of a concordance of differential glycopeptide abundance between severe inflammatory states (severe COVID-19 and sepsis) and controls is of interest in the context of a body of literature that documents glycoprotein sialylation and fucosylation in malignant disease and metastasis [25-27]. These observations are complemented by similar ones in inflammatory disease, where hypersialylation of immunoglobulins has been interpreted as representing an activated state of the immune system [28-30], which would certainly be consistent with the present context. While our data show similar patterns, larger within-group sample sizes need to be acquired to quantitatively and with sufficient statistical power assess associations of symptomatic COVID-19 and sepsis severity with both hyper-sialylation and hyper-fucosylation of glycopeptides.

Among 66 non-glycosylated peptides assayed, 25 were statistically significantly downregulated in both symptomatic COVID-19 and sepsis at FDR<0.05, while only seven were statistically significantly upregulated in both. While overall decreases of serum proteins have been reported in cancer in the past [31], a search of more recent literature to confirm this failed to yield additional evidence for this, and also not for inflammatory conditions. In a previous plasma proteomic analysis of ten sepsis patients, expression of APOC3 was statistically significantly downregulated the day after the suspected septic episode began compared with immediately after an elective surgical procedure [12]. While in our analysis the APOC3 peptide, GWVTDGFSSLK, was likewise somewhat decreased in septic patients compared to healthy controls, this difference did not reach statistical significance (fold change=0.956, FDR=0.903).

Our bioinformatic analysis highlighted a number of pathways that were statistically significantly altered among healthy subjects and COVID-19 patients, and among COVID-19 patients with either a symptomatic or asymptomatic disease course, specifically pathways involved with acute phase response signaling, the complement cascade, and the coagulation system. The changes in acute phase proteins can certainly be seen as reflecting immune responses to the viral infection, with the interleukin (IL)-6 cytokine family representing the statistically most significantly enriched upstream regulators among acute phase proteins identified as altered in the context of COVID-19. IL-6 is known to be a major regulator of acute phase protein synthesis, and IL-6 levels in serum have been reported to strongly correlate with COVID-19 infection [20] and risk of respiratory failure [21] in several studies.

The comparison between our study and the proteomic study conducted by Shen et al. [24] in a similar setting of severe and non-severe COVID-19 patients, indicating significant overlap of acute phase proteins that showed altered regulation among the 2 groups, supports the relevance of our findings. Meanwhile, the detection of a sizable number of additional altered acute phase glycoproteins indicates the importance of extending these analyses to include high-resolution characterization of post-translational modifications.

The activation of the complement cascade, illustrated in our study by the finding of a number of statistically significantly altered members of the pathway, including C3, C5, and C6, plays a key role in mediating immune response to viral infection and in promoting inflammatory processes through production of proinflammatory molecules. Using mice deficient in C3 (C3-/-), Gralinski et al. [22] evaluated complement activation in SARS-CoV-2 infection, and their results suggested that complement activation was involved in the pulmonary pathology and disease severity of SARS-CoV-2. Gao et al. [23] also reported increased protein levels of C5a in a small cohort of patients with severe COVID-19 disease.

The findings of the current analysis must be interpreted with some caution, primarily due to the limited number of samples available in each group, the opportunistic nature of gaining access to the samples that resulted in a paucity of more detailed annotations regarding demographic and clinical variables, such as patient ancestry, comorbidities, disease course, and ultimate outcome, and even complete information on age and sex. Moreover, we had to contend with both serum and plasma samples necessitating an inherently imperfect mathematical adjustment to normalize values. Also, while the blood samples of the severely ill COVID-19 patients were obtained upon presentation to their health care provider when they were acutely infected, prior to raising an immune response, the samples of subjects who had had an asymptomatic course of the illness were procured presumably after their infection had run its course, rendering them seropositive. While all these limitations are acknowledged, there is, however, a positive aspect: the many uncontrolled-for covariates that all these shortcomings introduced would be expected to dilute any between-group differences due to the resulting noise. Thus, the fact that, despite this, we found highly statistically significant results actually emphasizes the validity of our results, and the power of glycoproteomics, as the signals were strong enough to rise over all this noise. Our study draws additional indirect validation from the fact that we find the expected preponderance of men and older age groups among the patients who developed a severe case of the illness.

## Data Availability

All data produced in the present study are available upon reasonable request to the authors

## Author Contributions

Conceptualization, K.L.; methodology, G.X., R.R, and X.C.; software, D.S.; validation, K.L. and C.P.; formal analysis, C.P., P.R., and D. S., investigation, K.L., C.P.; resources, K.L., T.W., F. G., T. P., J. S, S. C., and K.E.; data curation, C.P. and P.R.; writing—original draft preparation, C.P., B.Z., K.L.; writing—review and editing, K.L., C.P., B.Z.,T.W., F.G., S.R.; visualization, B.Z. and C.P.; supervision, K.L.; project administration, K.L.; funding acquisition, K.L. All authors have read and agreed to the published version of the manuscript.

## Funding

This research received funding from InterVenn Biosciences. TTW was supported by Stanford University, the Chan Zuckerberg Biohub, the Searle Scholars Program, Fast Grants and the CEND COVID Catalyst Fund. Additional funding for TTW was received from the National Institute of Allergy and Infectious Diseases of the National Institutes of Health under award numbers U19AI111825 and the Stanford sample bank was supported in part by grant # UL1 TR001866 from the National Center for Advancing Translational Sciences (NCATS), National Institutes of Health (NIH) Clinical and Translational Science Award (CTSA) program.

## Institutional Review Board Statement

Ethical review and approval were waived for this study, due to use of fully de-identified samples, in accordance with the Common Rule (https://www.hhs.gov/ohrp/regulations-and-policy/regulations/finalized-revisions-common-rule/index.html).

## Informed Consent Statement

Patient consent was waived due to use of fully de-identified samples, in accordance with the Common Rule (https://www.hhs.gov/ohrp/regulations-and-policy/regulations/finalized-revisions-common-rule/index.html).

## Acknowledgments

none

## Conflicts of Interest

C.P., B.Z., P.R., G.X., R.R., K.L., D.S., and X.C. are full-time employees of InterVenn Biosciences which funded the study.

## Notes

### Competing Interest Statement

The authors have declared no competing interest.

### Funding Statement

This study was funded by InterVenn

### Author Declarations

Use of the samples for this study was approved by the Institutional Review Board of Stanford University under Study Review Protocol no. 55718.

